# Automated airway quantification associates with mortality in idiopathic pulmonary fibrosis

**DOI:** 10.1101/2023.03.29.23287913

**Authors:** Wing Keung Cheung, Ashkan Pakzad, Nesrin Mogulkoc, Sarah Needleman, Bojidar Rangelov, Eyjolfur Gudmundsson, An Zhao, Mariam Abbas, Davina McLaverty, Dimitrios Asimakopoulos, Robert Chapman, Recep Savas, Sam M Janes, Yipeng Hu, Daniel C. Alexander, John R Hurst, Joseph Jacob

**Affiliations:** Centre for Medical Image Computing, University College London, London, UK; Department of Computer Science, University College London, London, UK; Department of Medical Physics and Biomedical Engineering, University College London, London, UK; Department of Respiratory Medicine, Ege University Hospital, Izmir, Turkey; Medical School, University College London, London, UK; School of Clinical Medicine, University of Cambridge, Cambridge, UK; Interstitial Lung Disease Service, Department of Respiratory Medicine, University College, London Hospitals NHS Foundation Trust, London, UK; Department of Radiology, Ege University Hospital, Izmir, Turkey; Lungs for Living Research Centre, UCL, London, UK; UCL Respiratory, University College London, London, UK; Respiratory Medicine, Royal Free London NHS Foundation Trust, London, UK

## Abstract

**Objectives:** The study was to examine whether the airway metrics associate with mortality in IPF patients.

**Methods:** We performed an observational cohort study (n=90) of IPF patients identified from Ege University Hospital. An airway analysis tool AirQuant calculated median airway segmental intertapering and segmental tortuosity across 2^nd^ to 6^th^ generations of IPF airways. Intertapering measures the difference in median diameter between adjacent airway segments. Tortuosity evaluates the ratio of measured segmental length against direct end-to-end segmental length. Univariable linear regression analyses examined relationships between AirQuant variables, clinical variables and lung function tests. Univariable and Multivariable Cox proportional hazards models estimated mortality risk with the latter adjusted for patient age, gender, smoking status, antifibrotic use, CT usual interstitial pneumonia (UIP) pattern and either forced vital capacity (FVC) or diffusion capacity of carbon monoxide (DLco) if obtained within 3 months of the CT.

**Results:** No significant collinearity existed between AirQuant variables and clinical or functional variables. On univariable Cox regression analyses, male gender, smoking history, no antifibrotic use, reduced DLco, reduced segmental intertapering and increased segmental tortuosity associated with increased risk of death. On multivariable Cox regression analyses (adjusted using FVC), segmental intertapering (Hazard Ratio (HR)=0.75, 95% CI=0.66-0.85, p<0.001) and segmental tortuosity (HR=1.74, 95% CI=1.22-2.47, p=0.002) independently associated with mortality. Results were maintained with adjustment using DLco.

**Conclusions:** AirQuant generated measures of segmental intertapering and tortuosity independently associate with mortality in IPF patients. Abnormalities in proximal airway generations, which are not typically considered to be abnormal in IPF, have prognostic value.

**Key Points:**

- AirQuant generates measures of segmental intertapering and tortuosity.
- Automated airway quantification associates with mortality in IPF independent of established measures of disease severity.
- Automated airway analysis could be used to refine patient selection for therapeutic trials in IPF.

## Introduction

Idiopathic pulmonary fibrosis (IPF) is a chronic progressive fibrosing lung disease diagnosed using computed tomography imaging of the lungs. A hallmark of IPF is the presence on CT of honeycomb cysts and traction bronchiectasis in a subpleural, basal lower-zone predominant distribution ^1^. Traction bronchiectasis represents the pulling apart of airways walls by fibrotic contraction of the adjacent interstitial compartment of the lung. When honeycombing and traction bronchiectasis coexist in a subpleural, basal lower-zone predominant distribution ^1^, a patient can be ascribed a pattern of usual interstitial pneumonia (UIP). When traction bronchiectasis with the same distribution occurs in the absence of honeycomb cysts, a patient is ascribed a pattern of probable usual interstitial pneumonia (UIP). Post-hoc analyses of the INPULSIS trials and other studies have shown that a UIP pattern and a probable UIP pattern have similar associations with mortality, highlighting the prognostic importance of appropriately distributed traction bronchiectasis on CT.

Over the past 15 years numerous studies have examined the relationship between IPF imaging biomarkers and patient survival. Whilst these initially focussed on visual CT analysis, recent advances in computational image analysis including machine learning and deep learning have leveraged the volumetric nature of modern IPF imaging to consider three-dimensional imaging biomarkers. Vessel-related structures comprising pulmonary arteries, veins and associated fibrosis have been shown to associate strongly with mortality in patients across a variety of fibrosing lung diseases. Given the prognostic importance of traction bronchiectasis when scored visually, it is likely that computational measurement of airway abnormality in IPF might show promise as a prognostic biomarker.

Computational airway analysis on CT imaging is a two-stage process. The first stage constitutes segmentation of the airways of the lungs. Over the past 4 years, deep learning tools have been increasingly employed for airway segmentation, though in the main these tools have been trained to segment airways in patients with chronic obstructive pulmonary disease (COPD) rather than lung fibrosis. Nevertheless, tools trained on non-fibrosing lung diseases can be transferred to fibrosis scans and several proprietary and open-source tools are currently available for the acquisition of an airway segmentation of the lungs. Segmentation tools can show variable performance, with an acceptable standard of segmentation being the delineation of the first six generations of lung airways. We developed an in-house airway segmentation tool using a dilated 2D UNET deep learning framework which we used as input to our airway quantification pipeline.

The second stage of computational airway analysis comprises evaluation of the skeletonised airway segmentation. We have developed a computational airway tool called AirQuant which automatically identifies airway branching points and classifies the airway length between branching points into segments and hierarchical airway generations on a lobar basis. Using this framework AirQuant quantifies several airway metrics including the tapering of airway segments, segmental tortuosity and total segment count. AirQuant exists independent of airway segmentations. Accordingly in this study we examined whether AirQuant metrics for airway generations 2-6 associated with mortality in IPF patients.

## Materials and methods

In this observational cohort study, patients with a multidisciplinary team diagnosis of IPF and a volumetric inspiratory CT (slice thickness <1.25mm) were identified from Ege University Hospital, Izmir, Turkey (IPF diagnoses between 2008 and 2015). Clinical information obtained included patient age at time of CT acquisition, gender, smoking status (never versus ever), antifibrotic use (never versus ever), forced vital capacity (FVC) and diffusion capacity of carbon monoxide (DLco) obtained within 3 months of the CT, patient survival status and follow up time.

CT exclusion criteria included: CT slice thickness >1.25mm, radiologic evidence of lung infection, lung cancer, and/or likely acute exacerbation on CT and CT scan quality precluding visual assessment. Patients were excluded if they had under 3 months of follow-up following the baseline CT. Computational exclusion criteria included: poor airway segmentation quality, segmentation errors following use of the pulmonary toolkit and lobe classification errors (Figure 1). Approval for this retrospective study of clinically indicated pulmonary function and CT data was obtained from the local research ethics committees and Leeds East Research Ethics Committee: 20/YH/0120. The STROBE guidelines were adhered to in the design and analysis of this study (see STROBE checklist in online supplement).

**Figure 1:**
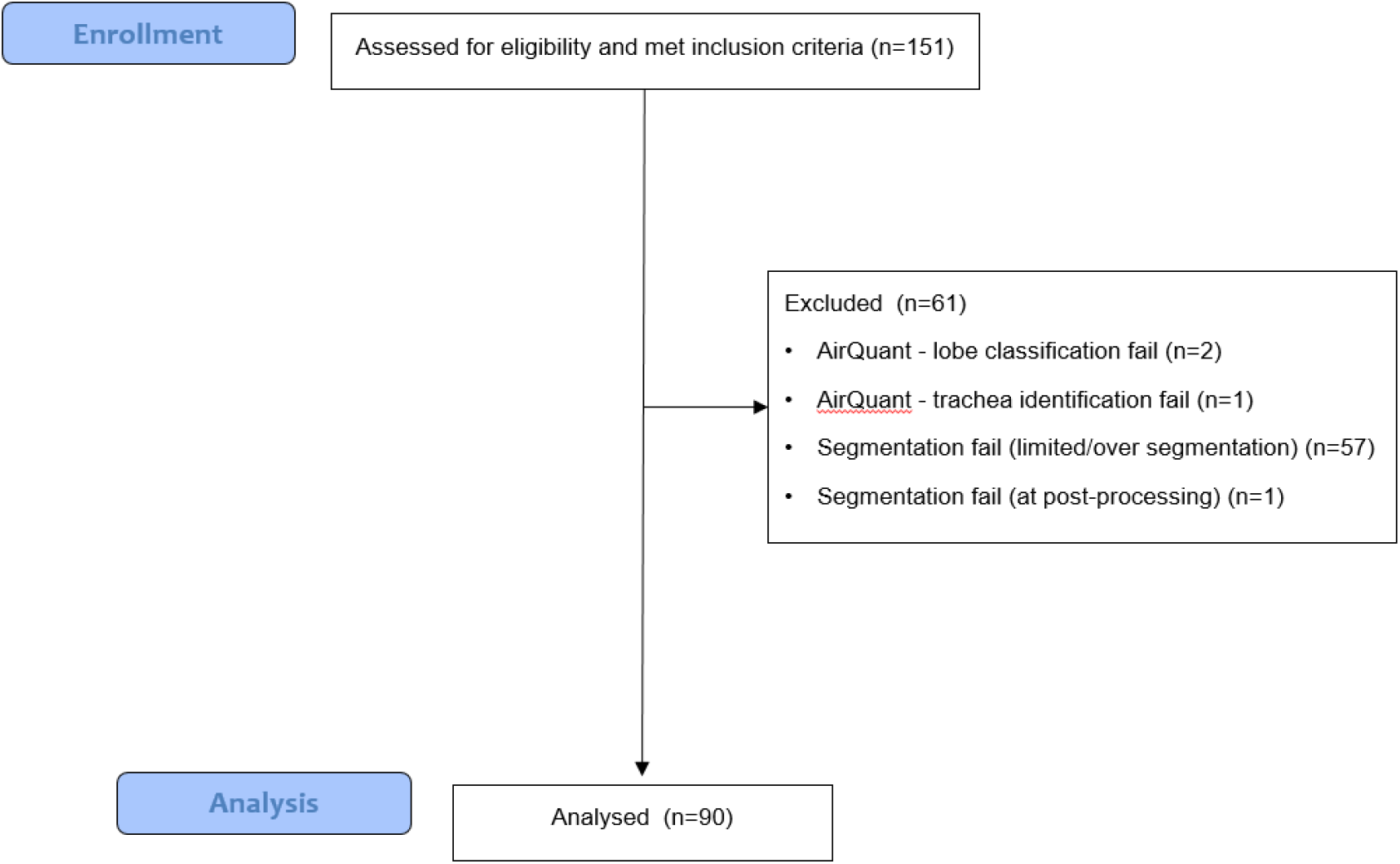
CONSORT diagram showing patient exclusions for the IPF study cohort.

### Visual CT Evaluation

A subspecialist radiologist (JJ) with 15-years thoracic imaging experience determined lobar percentages of interstitial lung disease (ILD) (sum of ground glass density, reticulation, traction bronchiectasis volume and honeycomb cysts, averaged across six lobes ^2^) and the presence of usual interstitial pneumonia (UIP) criteria^1^ (definite, probable, indeterminate) on baseline CTs. All patients with an indeterminate UIP pattern had histopathological confirmation of UIP.

### Computer-Based CT Evaluation

Details of the computational airway analysis pipeline^3^ have been previously described. Essentially this comprises two distinct stages: an airway segmentation for which we developed a novel algorithm described below, and AirQuant an airway analytic pipeline which exists independent of any airway segmentation.

#### Airway segmentation

Separate airway segmentations were performed using the pulmonary toolkit^4^ (PTK) software and a 2D dilated U-NET developed in-house. The 2D U-NET model was trained using 25 manually segmented airway trees from CTs in healthy subjects and patients with IPF. None of the CTs used to train the U-NET model were used in the current study. The airway segmentations produced by the pulmonary toolkit and 2D dilated U-NET were combined. A morphological closing procedure was then applied to re-connect airway segments that were disconnected following segmentation. The final airway tree mask was obtained by performing a largest connected components procedure.

#### Airway quantification using AirQuant

The key aim of this study was to evaluate the performance of AirQuant, an inhouse airway quantification pipeline. The technical details have been previously described^3^ and are briefly summarised here. CTs <1.25mm were selected for analysis and a standardised window level of -500 HU and window width of 1500 HU was used for viewing. AirQuant^3^ was used to calculate airway lumen diameter and length and measurements derived from these, across all segmented airways on a CT scan. AirQuant works in a series of stages. First, an acyclic skeleton is derived from an airway segmentation by propagating a splitting wave from the trachea^4^ and applying thinning to the airways to derive an airway skeleton^5^. Second, the skeleton is converted into a graph, separating airway segments into individual components. An airway segment is defined as a single branch between splitting or endpoints. Third, airway segments are automatically classified into their lung lobes, with the lingula interpreted as the left middle lobe. A new airway generation is produced every time an airway divides and airway generations are counted sequentially from the trachea. Airway segments are classified according to their lobar generation. Fourth, polynomial splines are used to facilitate interpolation along the branch length so that measurements can be made at intervals. This interval is dynamically set to half the size of the smallest voxel dimension. Fifth, perpendicular airway CT slices are derived at the chosen spline interval. The size of these slices are a maximum of 40mm, enough to frame the airway boundaries. Finally, measurements are made on these interpolated slices using the Full Width at Half Maximum edgecued segmentation limited technique^5^. An intensity profile is taken from the center of the airway outwards at multiple angles, and the airway wall is modelled as a gaussian curve. The inner edge of the gaussian curve is determined to be the inner boundary of the airway lumen. An ellipse is then fitted to these boundary points. The diameter used to represent the airway at that point is computed by 2x radius, where the radius represents the square root of the product of the minor and major axis radii.

Modelling the airways in this way allows systematic analysis of the airways. AirQuant variables evaluated in the study included: segmental intertapering, segmental tortuosity, and total segment count. Segmental intertapering between adjacent segments represented the mean diameter of an airway segment subtracted from the mean diameter of the parent airway segment, with the result divided by the mean diameter of the parent airway segment (Figure 2). As airways do not taper/narrow in diameter as expected when extending into fibrotic lung, airway segmental intertapering/narrowing would be expected to reduce. Measures of segmental tortuosity rely on calculating the Euclidean length of an airway segment. Euclidean length is determined by the arc-length between the start and end points of the airway spline (Figure 2). The ratio of arc-length to Euclidean length (Figure 2) denotes the degree of tortuosity of that segment, which we expect to be lower in healthy individuals. Total segment count represents the total number of airway segments identified on a CT. 3D plots of the airway tree of an IPF patient are shown in Figure 3 and the corresponding airway graph representation is shown in Figure 4.

**Figure 2:**
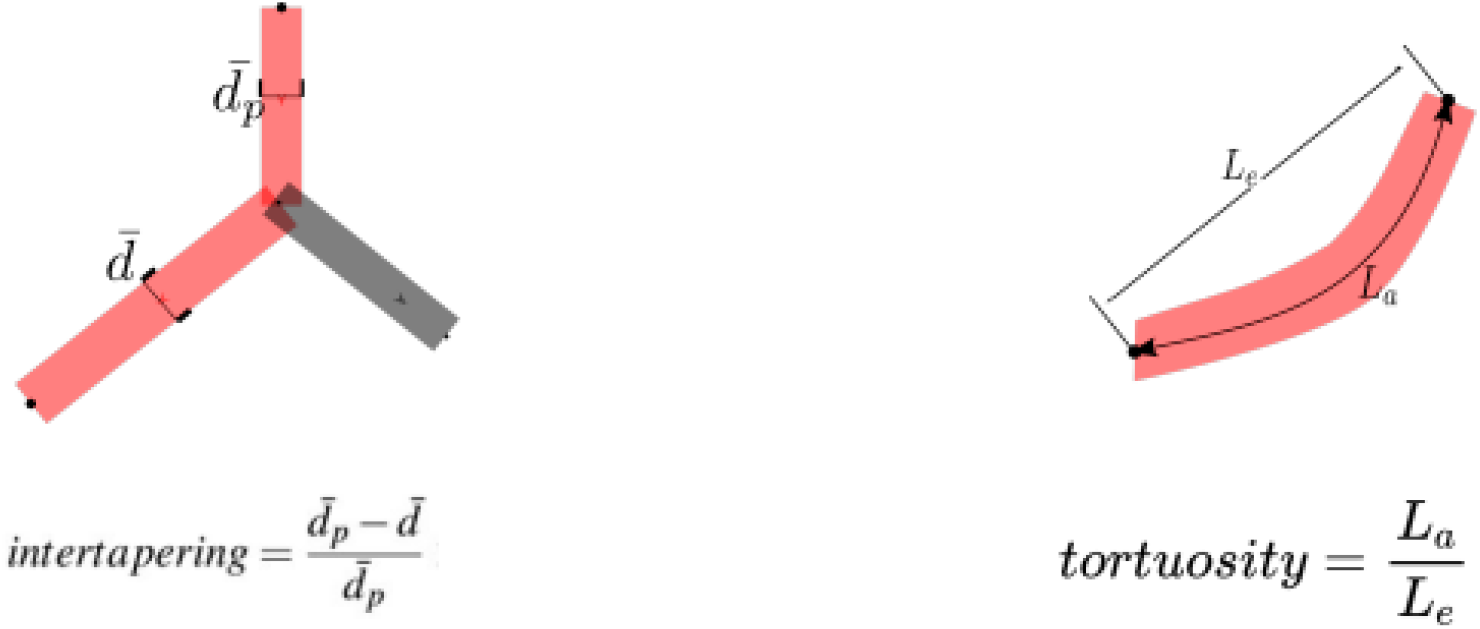
A schematic diagram illustrating the calculations used to derive segmental intertapering and segmental tortuosity.

**Figure 3:**
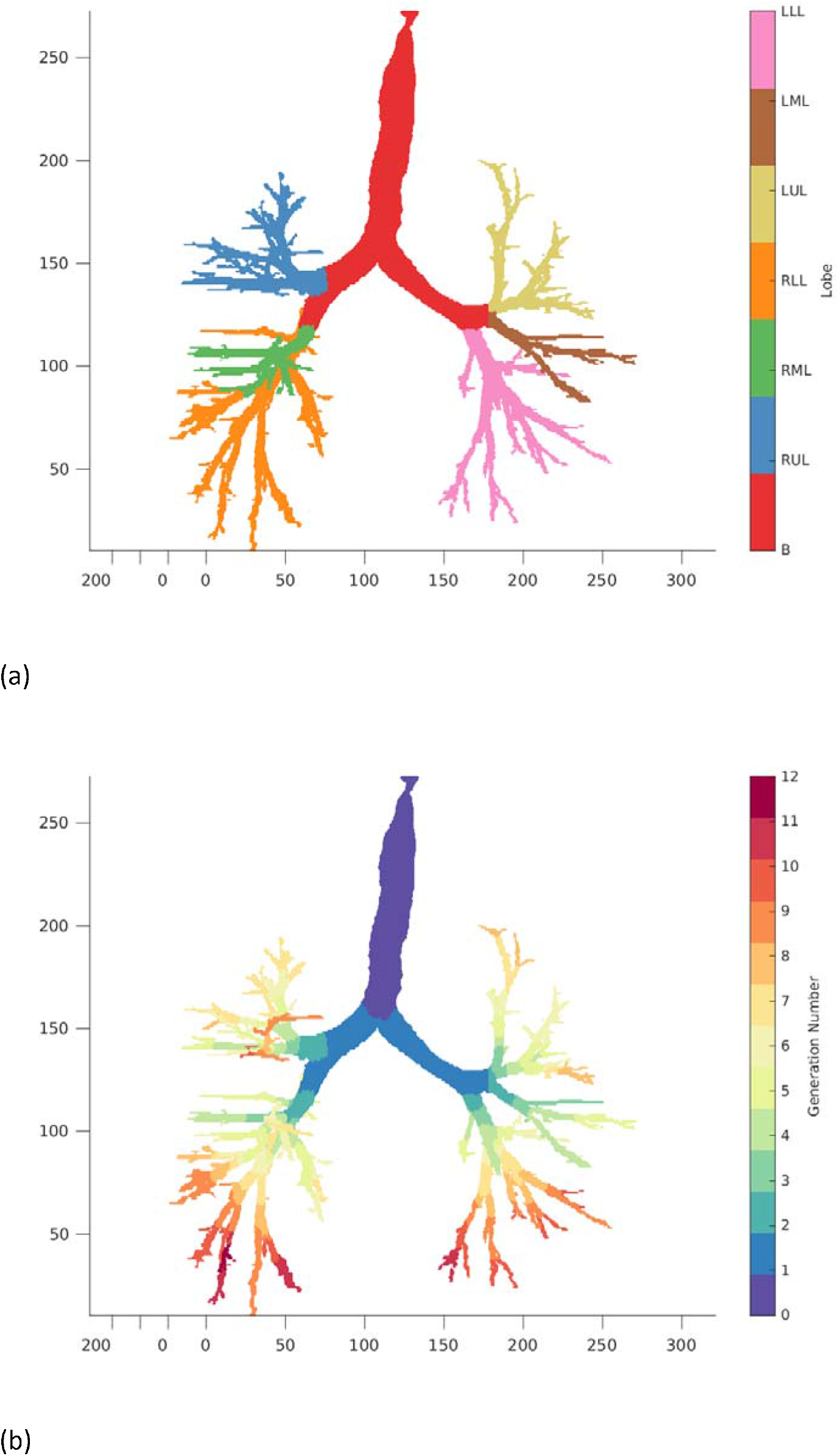
Example of an IPF patient (age: 77.9, gender: female, baseline FVC: 120%, DLco: 57%, UIP category: definite and time to death: 5.06 years) derived from volumetric computed tomography imaging using AirQuant. (a) 3D airway with lobe labels (b) 3D airway with generation labels

**Figure 4:**
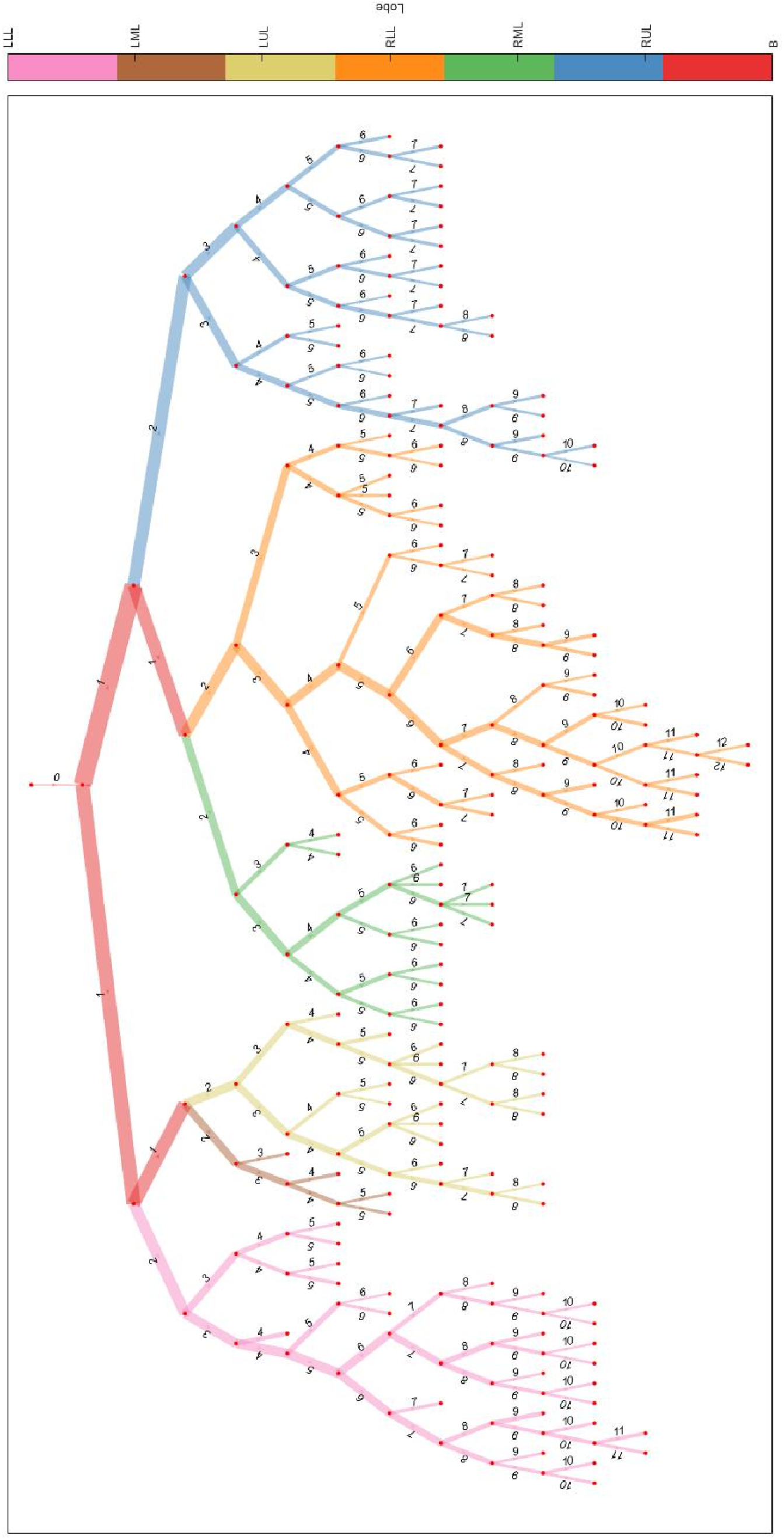
Airway graph representation of airway divisions are represented by nodes and airway segments by edges. Edge thickness is proportional to average luminal diameter. The edge label represents the lobar generation of each airway segment: 0 for the trachea; 1 for the main and intermediate bronchi (B); 2 for the first intralobar airway; 3 onwards for each subsequent airway division. Edge colour is coded to lobe classification, RUL=right upper lobe, LUL=left upper lobe, RML=right middle lobe, LML=left middle lobe, RLL=right lower lobe, LLL=left lower lobe.

The analysis considered airways in generation 2-6 in the lungs, analysed on a lobar basis. It aimed to evaluate AirQuant performance on airway segmentations trees that would be achievable by the majority of good quality airway segmentation tools currently available to researchers. We also specifically wanted to examine whether changes in proximal airways, which are not traditionally thought to be abnormal in IPF could provide an association with mortality.

### Statistical analysis

Data are presented as patient proportions (percentages) or means (with standard deviations) or medians (with range of values), as appropriate. Univariable linear regression analyses were performed to examine relationships between AirQuant variables and clinical variables, lung function tests and visual CT ILD extent. Univariable and Multivariable Cox proportional hazards models were performed with the latter adjusted for patient age, gender, smoking status, antifibrotic use and either FVC or DLco. Cox regression models were investigated for proportionality using plots of scaled Schoenfeld residuals. A p-value <0.05 was considered significant across all analyses. Descriptive statistics, linear regression and Cox regression analyses were performed on SPSS (version 27, IBM, New York, USA).

## Results

Demographic data, baseline FVC and DLco values, and visual and AirQuant CT measures for IPF cohort (n=90) is shown in Table 1. The mean patient age was 66 years with a median follow up time of 2.7 years. Mean baseline FVC was 77.2% and mean baseline DLco was 50.6%.

**Table 1:**
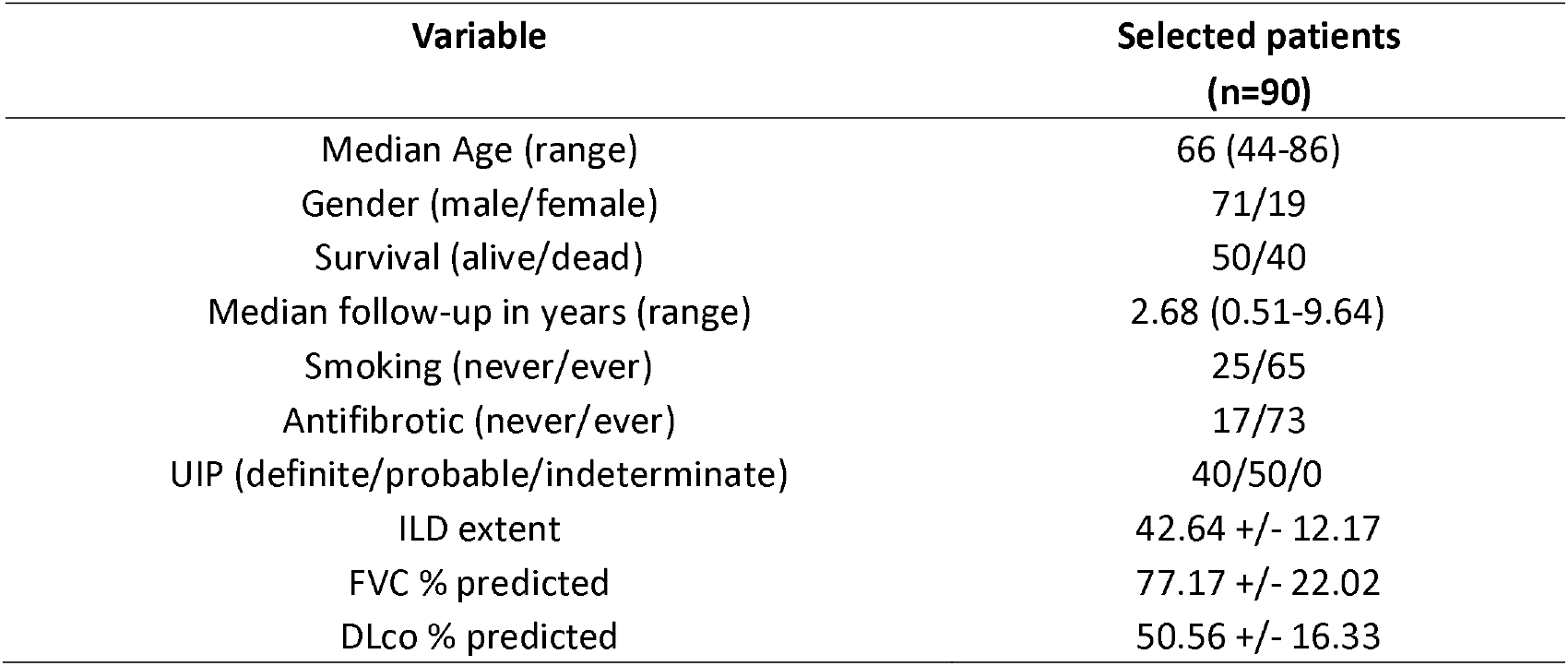
Patient demographics and pulmonary function indices in selected patients of IPF patients are described as mean and standard deviations, except where noted. UIP = usual interstitial pneumonia, ILD = Interstitial lung disease, FVC = forced vital capacity, DLco = diffusing capacity for carbon monoxide.

| Variable | Selected patients<br>(n=90) |
| --- | --- |
| Median Age (range) | 66 (44-86) |
| Gender (male/female) | 71/19 |
| Survival (alive/dead) | 50/40 |
| Median follow-up in years (range) | 2.68 (0.51-9.64) |
| Smoking (never/ever) | 25/65 |
| Antifibrotic (never/ever) | 17/73 |
| UIP (definite/probable/indeterminate) | 40/50/0 |
| ILD extent | 42.64 +/- 12.17 |
| FVC % predicted | 77.17 +/- 22.02 |
| DLco % predicted | 50.56 +/- 16.33 |

Univariable linear regression showed no significant collinearity between AirQuant variables and patient age, gender, smoking status, antifibrotic use, visual ILD extent, baseline FVC and DLco. On univariable Cox regression analyses, male gender, a history of smoking, no antifibrotic use, reduced DLco, reduced segmental intertapering and increased segmental tortuosity associated with increased risk of death.

On multivariable Cox regression analyses considering CT UIP pattern and disease severity adjustment using FVC, segmental intertapering (Hazard Ratio (HR)=0.75, 95% CI=0.66-0.85, p<0.001) and segmental tortuosity (HR=1.74, 95% CI=1.22-2.47, p=0.002) independently associated with mortality (Table 2). Results were maintained with separate adjustment using DLco (Table 3) and visual ILD extent (data not shown).

**Table 2:** Multivariable Cox regression models showing mortality in IPF cohorts (n=90) using segmental intertapering and segment tortuosity in airway generations 2-6. Models were adjusted for patient age, gender, smoking, antifibrotic, usual interstitial pneumonia (UIP) and forced vital capacity (FVC).

| Variable | Hazard ratio | 95% CI-lower | 95% CI-upper | P value |
| --- | --- | --- | --- | --- |
| Age | 1.003 | 0.955 | 1.054 | 0.901 |
| Gender | 0.244 | 0.051 | 1.154 | 0.075 |
| Smoking | 2.367 | 0.685 | 8.174 | 0.173 |
| Antifibrotic | 0.249 | 0.099 | 0.627 | 0.003 |
| UIP | 0.451 | 0.218 | 0.933 | 0.032 |
| FVC | 1.011 | 0.991 | 1.031 | 0.293 |
| Segmental intertapering | 0.749 | 0.664 | 0.845 | <0.001 |
| Age | 1.024 | 0.975 | 1.075 | 0.348 |
| Gender | 0.151 | 0.033 | 0.692 | 0.015 |
| Smoking | 1.527 | 0.468 | 4.988 | 0.483 |
| Antifibrotic | 0.273 | 0.109 | 0.686 | 0.006 |
| UIP | 0.575 | 0.269 | 1.226 | 0.152 |
| FVC | 1.007 | 0.988 | 1.027 | 0.459 |
| Segment tortuosity | 1.736 | 1.222 | 2.466 | 0.002 |

**Table 3:** Multivariable Cox regression models showing mortality in IPF cohorts (n=90) using segmental intertapering and segment tortuosity in airway generations 2-6. Models were adjusted for patient age, gender, smoking, antifibrotic, usual interstitial pneumonia (UIP) and diffusing capacity for carbon monoxide (DLco).

| Variable | Hazard ratio | 95% CI-lower | 95% CI-upper | P value |
| --- | --- | --- | --- | --- |
| Age | 1.026 | 0.974 | 1.082 | 0.328 |
| Gender | 0.599 | 0.097 | 3.703 | 0.581 |
| Smoking | 3.173 | 0.738 | 13.643 | 0.121 |
| Antifibrotic | 0.260 | 0.093 | 0.728 | 0.010 |
| UIP | 0.598 | 0.262 | 1.367 | 0.223 |
| DLco | 0.974 | 0.941 | 1.008 | 0.133 |
| Segmental intertapering | 0.769 | 0.670 | 0.882 | <0.001 |
| Age | 1.047 | 0.992 | 1.105 | 0.097 |
| Gender | 0.297 | 0.055 | 1.608 | 0.159 |
| Smoking | 2.095 | 0.523 | 8.393 | 0.296 |
| Antifibrotic | 0.316 | 0.115 | 0.868 | 0.025 |
| UIP | 0.755 | 0.318 | 1.794 | 0.525 |
| DLco | 0.970 | 0.939 | 1.002 | 0.069 |
| Segment tortuosity | 1.818 | 1.204 | 2.746 | 0.004 |

## Discussion

Our pilot study has highlighted the potential for automated airway quantification, specifically segmental intertapering and segmental tortuosity to be used as a prognostic tool in the assessment of patients with IPF. The two AirQuant metrics associated with mortality irrespective of the baseline severity of disease (as measured by FVC or DLco or visual ILD extent) and regardless of the type of UIP pattern seen on the CT.

Our observation that the proximal airways are abnormal in IPF confirm results from previous studies that have shown increased volumes^11^ and reduced resistance of conducting airways in IPF^12^ patients. Studies using aerosol-derived morphometry have also shown that increased airway dimensions are visible throughout the airway tree of IPF lungs^13^, mirroring our finding of increased proximal airway tortuosity. It is possible that proximal airway dilatation represents a potential surrogate for morphologically extensive interstitial damage on CT, given the correlations between visually scored ILD extent and segmental intertapering and tortuosity. However, it is notable that functional measures of disease severity (FVC and DLco) only correlated weakly with AirQuant metrics of segmental intertapering and tortuosity.

One of the advantages of CT imaging over lung function measurements lies in the ability of CT to provide localised estimations of damage, whilst lung function provides averaged global measures of lung disease. Existing quantitative tools applied to the lungs have primarily quantified lung damage at a global level^8^ or examined damage within lung zones^9^ (upper, middle and lower). AirQuant metrics however provide a more granular estimate of lung damage by assessing airways at a generational level. To aid interpretation of AirQuant, airway maps were developed that provide an intuitive visualisation of regions of airway damage.

A UIP pattern has been shown in numerous prior studies to be a strong prognostic indicator in patients with IPF^10^. Yet in our study AirQuant measures of segmental intertapering and tortuosity showed consistently stronger associations with mortality than a UIP pattern. In IPF patients, honeycombing may only constitute a small volume of the lung, and the presence of a UIP pattern therefore may not capture the extent of an individual’s disease. Disease extent may be better captured by identifying a reduction in mean tapering or an increase in tortuosity of all the airways in an entire generation in the lung.

There were limitations to the current study. Though the segmentation quality of our in-house algorithm is certainly comparable to tools being used commercially, the heterogeneity of CT acquisitions and imaging constraints such as breathing artefacts made following airway courses deep into the lungs challenging for the segmentation software in certain cases. This was accentuated in patients with more severe disease who as a result had CTs excluded more frequently. AirQuant itself rarely failed (failure seen in 2% of cases) if provided with an adequate segmentation and remained robust across a variety of CT acquisition parameters, though our results will need to be confirmed in validation populations. Though this was not a study of airway segmentation performance, our results suggest that airway segmentation tools are likely to perform better in patients with early IPF and may have value in cohort enrichment of therapeutic trials^14^.

In conclusion, our pilot study demonstrated that AirQuant generated measures of airway abnormality (segmental intertapering and tortuosity) significantly associate with mortality in patients with IPF. We also highlight that airway abnormalities in proximal airway generations which are not typically considered to be abnormal in IPF have prognostic value.

## Data Availability

All data produced in the present study are available upon reasonable request to the authors

## Abbreviations

CI: Confidence interval
COPD: Chronic obstructive pulmonary disease
CT: Computerised tomography
DLco: Diffusion capacity of carbon monoxide
FVC: Forced vital capacity
HR: Hazard ratio
HU: Hounsfield unit
ILD: Interstitial lung disease
IPF: Idiopathic pulmonary fibrosis
PTK: Pulmonary toolkit
SPSS: Statistical Product and Service Solutions
STROBE: The Strengthening the Reporting of Observational Studies in Epidemiology
UIP: Usual interstitial pneumonia

## Acknowledgments

This research was funded in whole or in part by the Wellcome Trust [209553/Z/17/Z]. For the purpose of open access, the author has applied a CC-BY public copyright licence to any author accepted manuscript version arising from this submission. This project, JJ, EG, ED, and SMJ were also supported by the NIHR UCLH Biomedical Research Centre, UK.

## Author Contributions

JJ, WKC and AP provided substantial contributions to the conception and design of the work, the acquisition, analysis, and interpretation of data for the work; AND drafted the work and revised it critically for important intellectual content; AND gave final approval of the version to be published; AND agree to be accountable for all aspects of the work in ensuring that questions related to the accuracy or integrity of any part of the work are appropriately investigated and resolved.

JRH, SN, BR, EG, AZ, MA, DM, DA, RC, RS, YH, DCA, NM, SMJ provided substantial contributions to the acquisition, analysis and interpretation of data for the work; AND revised it critically for important intellectual content; AND gave final approval of the version to be published; AND agree to be accountable for all aspects of the work in ensuring that questions related to the accuracy or integrity of any part of the work are appropriately investigated and resolved.

